# SARS-CoV-2 infections and antibody responses among health care workers in a Spanish hospital after a month of follow-up

**DOI:** 10.1101/2020.08.23.20180125

**Authors:** Gemma Moncunill, Alfredo Mayor, Rebeca Santano, Alfons Jiménez, Marta Vidal, Marta Tortajada, Sergi Sanz, Susana Méndez, Anna Llupià, Ruth Aguilar, Selena Alonso, Diana Barrios, Carlo Carolis, Pau Cisteró, Eugenia Chóliz, Angeline Cruz, Silvia Fochs, Chenjerai Jairoce, Jochen Hecht, Montserrat Lamoglia, Mikel J. Martínez, Javier Moreno, Robert A. Mitchell, Natalia Ortega, Nuria Pey, Laura Puyol, Marta Ribes, Neus Rosell, Patricia Sotomayor, Sara Torres, Sarah Williams, Sonia Barroso, Anna Vilella, Antoni Trilla, Pilar Varela, Carlota Dobaño, Alberto L Garcia-Basteiro

**Affiliations:** ISGlobal, Hospital Clínic - Universitat de Barcelona, Barcelona, Catalonia, Spain; Spanish Consortium for Research in Epidemiology and Public Health (CIBERESP), Madrid, Spain.; Centro de Investigação em Saúde de Manhiça (CISM), Maputo, Mozambique; Occupational Health Department, Hospital Clínic, Universitat de Barcelona, Barcelona, Spain.; Department of Basic Clinical Practice, Faculty of Medicine, Universitat de Barcelona, Spain; Department of Preventive Medicine and Epidemiology, Hospital Clinic, Universitat de Barcelona, Spain; Centre for Genomic Regulation (CRG), The Barcelona Institute of Science and Technology, Barcelona, Spain; Faculty of Medicine and Health Sciences, Universitat de Barcelona, Spain; Faculty of Health Sciences of Blanquerna. Universitat Ramon Llull de Barcelona.; Department of Microbiology, Hospital Clínic, Universitat de Barcelona, Spain; International Health Department, Hospital Clínic, Universitat de Barcelona, Spain

**Keywords:** COVID-19, SARS-CoV-2, seroprevalence, antibodies, health care workers, longitudinal cohort, kinetics

## Abstract

**Background:** At the peak of the COVID-19 pandemic in Spain, cumulative prevalence of SARS-CoV-2 infection in a cohort of 578 randomly selected health care workers (HCW) from Hospital Clínic de Barcelona was 11.2%.

**Methods:** A follow-up survey one month after the baseline (April-May 2020) measured SARS-CoV-2 infection by real time reverse-transcriptase polymerase chain reaction (rRT-PCR) and IgM, IgA, IgG and subclasses to the receptor-binding domain of the SARS-CoV-2 spike protein by Luminex. Prevalence of infection was defined by a positive SARS-CoV-2 rRT-PCR and/or antibody seropositivity.

**Results:** The cumulative prevalence of infection at month 1 was 14.9% (84/565) and the seroprevalence 14.5% (82/565) for IgM and/or IgG and/or IgA. We found 25 (5%) new infections in participants without previous evidence of infection at baseline (501) and two participants seroreverted for IgM and/or IgG and/or IgA. Among seropositive participants at baseline, IgM and IgA levels generally declined at month 1 (antibody decay rates of 0.49 (95% CI, 0.40-0.60) and 0.34 (95% CI, 0.26-0.44)), respectively. Eight percent of the participants seroreverted for IgM and 11% for IgA. Subjects reporting COVID-19-like symptoms and laboratory and other technicians had higher risk of infection. The most frequent subclass responses were IgG1 and IgG2, followed by IgG3, with higher levels of IgG1, and only IgA1 but no IgA2 was detected.

**Conclusions:** Our findings highlight the importance of a continuous and improved surveillance of SARS-CoV-2 infections in HCW, particularly in high risk groups. The decay of IgA and IgM levels have implications for seroprevalence studies using these isotypes.

## INTRODUCTION

Since the start of the coronavirus disease 2019 (COVID-19) pandemic, caused by severe acute respiratory syndrome coronavirus 2 (SARS-CoV-2), there have been two priority questions: to establish the prevalence and incidence of the infection and to unravel whether cases are protected from future reinfections and/or disease. Among the 14.4 million confirmed SARS-CoV-2 infections and 603,691 deaths, as of July 2020 (https://covid19.who.int/), health care workers (HCW) continue to be one of the populations at higher risk due to close contact with COVID-19 patients [1]. To date, it is estimated that more than 230,000 HCW have been infected and over 3000 have died of COVID-19 [2]. Nevertheless, most infections in HCW are asymptomatic or mild [1,3–6] but undetected infections can put their HCW fellows and patients at risk. Prompt identification of cases by real time reverse-transcriptase polymerase chain reaction (rRT-PCR) screenings at hospitals is crucial to avoid new infections, isolations and quarantines in HCW.

We previously reported the prevalence of SARS-CoV-2 in a cohort of 578 HCW from a large hospital from Barcelona, Spain, at the peak of the pandemic (baseline, March 28^th^ to April 9^th^, 2020) [3]. We found that 9.3% (95% CI: 7.1–12.0) of the participants were seropositive and the cumulative prevalence of SARS-CoV-2 infection (considering a past or current positive result to either antibody testing or rRT-PCR) was 11.2% (95% CI: 8.8–14.1). The seroprevalence was relatively low but higher than the 7% estimated in the general population in Barcelona one month later according to a large national seroprevalence study [7]. Our findings were consistent to other studies in HCW [4,5,8], although prevalence of up to 44% had also been reported in other countries [9]. Importantly, 40% of the infections in our HCW cohort had not been previously detected [3].

This cohort is being followed up over a year to assess seroconversion and to understand naturally acquired immunity to COVID-19 by evaluating the kinetics of antibody responses, including IgG subclasses that have barely been explored [10,11]. Each IgG subclass results in different antibody functions beyond viral neutralization through the differential binding of Fc receptors or to complement, therefore this characterization is relevant to understand the mechanisms of immune protection [12].

Here, we determined the prevalence of SARS-CoV-2 by antibody serology and rRT-PCR one month after the baseline. We measured IgM, IgG, and IgA isotypes and subclasses, and assessed the factors associated with new infections as well as levels and kinetics of antibodies.

## METHODS

### Study Design and Population

We performed the second cross-sectional survey (April 27^th^ to May 6^th^, 2020) of a 4-stage seroprevalence study in a cohort of 578 HCW who had been randomly selected and recruited from a total of 5598 HCW registered at Hospital Clínic de Barcelona (HCB) [3]. Participants were invited to a follow-up visit one month later. The study population included HCW who deliver care and services directly or indirectly to patients. Further information can be found in supplementary materials. We collected a nasopharyngeal swab for SARS-CoV-2 rRT-PCR and a blood sample for antibody and immunological assessments. For participants isolated at home due to a COVID-19 diagnosis or on quarantine, data and sample collection took place at their households. Written informed consent was obtained from all study participants prior to study initiation. The study was approved by the Ethics Committee at HCB (Ref number: HCB/2020/0336). Data for each participant was collected in a standardized electronic questionnaire as it has been previously described [3].

### SARS-CoV-2 Laboratory Analyses

Methods for SARS-CoV-2 detection by rRT-PCR have follow the CDC-006-00019 CDC/DDID/NCIRD/ Division of Viral Diseases protocol, as previously described.[3] (Supplementary information). IgM, IgG and IgA antibodies to receptor-binding domain (RBD) of the spike glycoprotein of SARS-CoV-2, kindly donated by the Krammer lab (Mount Sinai, New York) [13], were measured as in the baseline survey of this cohort [3]. IgG and IgA subclass assays were performed following a similar Luminex protocol (Supplementary Information) [14]. Assay cutoff was calculated as 10 to the mean plus 3 standard deviations of log_10_-transformed MFIs of 47 pre-pandemic controls.

### Statistical Analysis

We tested the association between variables with the Chi-square or Fisher’s exact test (for categorical variables), T Student or Wilcoxon Sum Rank tests (for continuous quantitative variables). Univariable logistic models were run to evaluate factors associated with seropositivity. The effect of infection on antibody levels was analyzed using multilevel mixed-effects linear regression models incorporating Gaussian random intercepts. This resulted in an estimate of the rates of antibody dynamics (decay), assuming a single exponential model. Cumulative seropositive data was generated by selecting antibody levels at month 0 from individuals who were seropositive and antibody levels from month 1 in individuals who seroconverted from month 0 to month 1 and used for the analysis of antibody levels by different factors. The LOESS (locally estimated scatterplot smoothing) method was used to fit a curve to depict kinetics of antibody levels over time. Statistical comparisons were performed at two-sided significance level of 0.05 and 95% confidence intervals (CI) were calculated for all estimations. Analyses were undertaken using Stata/SE software version 16.1 and R studio version R-3.5.1 [15] (packages tidyverse and pheatmap).

## RESULTS

### Demographic Characteristics of Individuals without Previous Evidence of Infection

One month after baseline, 566 out of 578 HCW were visited again (2.1% of lost to follow-up) and blood sample was obtained in 565 of them. From the total of 566 individuals, 65 (11.5%) had previous evidence of infection at baseline by serology or rRT-PCR [3], thus the remaining 501 (88.5%) individuals had no evidence of infection. Of those, 359 (71.7%) were female, 268 (53.5%) were younger than 45 years of age and the mean age was 42 years. Half of the individuals (239/501) were nurses, auxiliary nurses or stretcher-bearers, 25.5% (128/501) were physicians and 38/501 7.6% were lab or other technicians; 50.3% (252/501) worked in COVID-19 units and 76.4% (383/501) had direct contact with COVID-19 patients since the last visit. Eleven per cent (54/501) participants reported having had COVID-19-compatible symptoms in the previous month and 21.2% (106/501) had co-morbidities (Supplementary table 1).

### SARS-CoV-2 Infections in a Month of Follow-up

At month 1 visit, the cumulative prevalence of infection measured by either rRT-PCR or serology was 14.9% (84/565). Nine participants had a positive rRT-PCR in the28 days following the initial study visit, and only 3 of these were detected at the second survey. The seroprevalence at month 1 was 14.5% (82/565) for either IgM and/or IgG and/or IgA and 10.1% for IgM, 11.3% for IgG and 11.5% for IgA (Supplementary Figure 1). There was an absolute increment of 25 SARS-CoV-2 infections detected by rRT-PCR or serology, 5% among the 501 previously uninfected individuals (4% from all individuals at month 1). Among these 25 individuals, infection was detected only by antibody serology (IgM/IgG/IgA) in 16, by antibody serology and rRT-PCR in 7, and only by rRT-PCR in 2. The latter two seronegative individuals at month 1 had the positive rRT-PCR result more than 20 days before the survey.

Having had COVID-19 compatible symptoms during the follow-up month was associated with experiencing a SARS-CoV-2 infection between month 0 and 1 with an OR of 6.55 (95% CI 2.77-14.44) and a p< 0.0001 in univariable analysis (Table 1). Professional category was also associated with infections; lab and other technicians had higher odds of being infected (OR 13.3; 95% CI 1.47-115.76; p = 0.048). Sex, age, co-morbidities, working in COVID-19 units, or having had direct contact with patients since the last visit were not associated with SARS-CoV-2 infection.

**Table 1.**
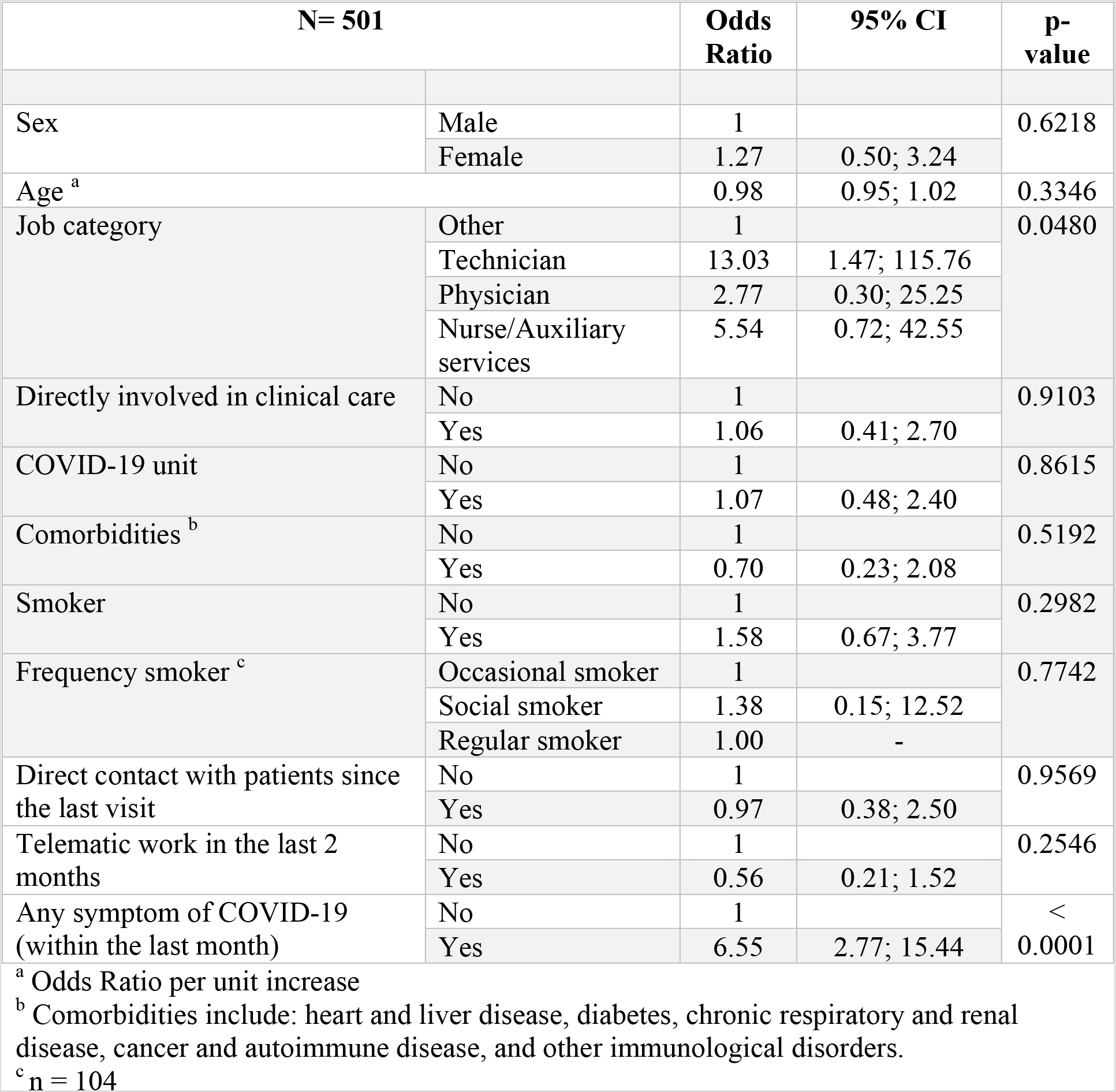
Factors associated with SARS-CoV-2 infections from recruitment to month 1

| N= 501 |  | Odds Ratio | 95% CI | p-value |
| --- | --- | --- | --- | --- |
| Sex | Male | 1 |  | 0.6218 |
|  | Female | 1.27 | 0.50; 3.24 |  |
| Age <sup>a</sup> |  | 0.98 | 0.95; 1.02 | 0.3346 |
| Job category | Other | 1 |  | 0.0480 |
|  | Technician | 13.03 | 1.47; 115.76 |  |
|  | Physician | 2.77 | 0.30; 25.25 |  |
|  | Nurse/Auxiliary services | 5.54 | 0.72; 42.55 |  |
| Directly involved in clinical care | No | 1 |  | 0.9103 |
|  | Yes | 1.06 | 0.41; 2.70 |  |
| COVID-19 unit | No | 1 |  | 0.8615 |
|  | Yes | 1.07 | 0.48; 2.40 |  |
| Comorbidities <sup>b</sup> | No | 1 |  | 0.5192 |
|  | Yes | 0.70 | 0.23; 2.08 |  |
| Smoker | No | 1 |  | 0.2982 |
|  | Yes | 1.58 | 0.67; 3.77 |  |
| Frequency smoker <sup>c</sup> | Occasional smoker | 1 |  | 0.7742 |
|  | Social smoker | 1.38 | 0.15; 12.52 |  |
|  | Regular smoker | 1.00 | - |  |
| Direct contact with patients since the last visit | No | 1 |  | 0.9569 |
|  | Yes | 0.97 | 0.38; 2.50 |  |
| Telematic work in the last 2 months | No | 1 |  | 0.2546 |
|  | Yes | 0.56 | 0.21; 1.52 |  |
| Any symptom of COVID-19 (within the last month) | No | 1 |  | < 0.0001 |
|  | Yes | 6.55 | 2.77; 15.44 |  |

<sup>a</sup> Odds Ratio per unit increase<sup>b</sup> Comorbidities include: heart and liver disease, diabetes, chronic respiratory and renal disease, cancer and autoimmune disease, and other immunological disorders.<sup>c</sup> n = 104

### SARS-CoV-2 Antibody Seroreversion Rate

From the 54 seropositive HCW at baseline, 1 did not have sample available at month 1, 3/36 (8.33%) seroreverted for IgM, 1/44 (2.3%) for IgG and 5/47 (10.6%) for IgA. From the remaining 512 HCW who were seronegative at recruitment, a total of 31 HCW (6.1%) seroconverted during the follow-up month for at least one immunoglobulin. Of those, 8 had a positive rRT-PCR detected at baseline or before. Separately by isotypes, 21 seroconverted for IgM, 20 for IgG and 23 for IgA. There were 5 individuals who seroconverted for IgM only, 3 for IgG only, and 6 for IgA only.

There were 5 seronegative HCWs at month 1 but with a previous positive rRT-PCR. Time since the first positive rRT-PCR ranged from 20 to 47 days. Two of these HCW were asymptomatic.

### IgA, IgG and IgM Levels in Seropositive HCW

Overall, IgM and IgA levels decreased from baseline to month 1, with antibody decay rates of 0.49 (95% CI, 0.40-0.60) and 0.34 (95% CI, 0.26-0.44), respectively (Table 2). The estimated time to seroreversion was 2.77 months (95% CI, 2.16-3.83, p-value < 0.0001) and 2.26 months (1.81-2.99, p-value < 0.0001) for IgM and IgA, respectively. In contrast, overall IgG levels did not change between timepoints (Table 2), however, they decreased in 14/43 individuals and increased in 28/43 individuals (Figure 1). In adjusted models by days since onset of symptoms, antibody decay rates were similar (Supplementary Table 2). No differences in antibody kinetics were observed between asymptomatic and symptomatic individuals (Figure 1).

**Table 2.**
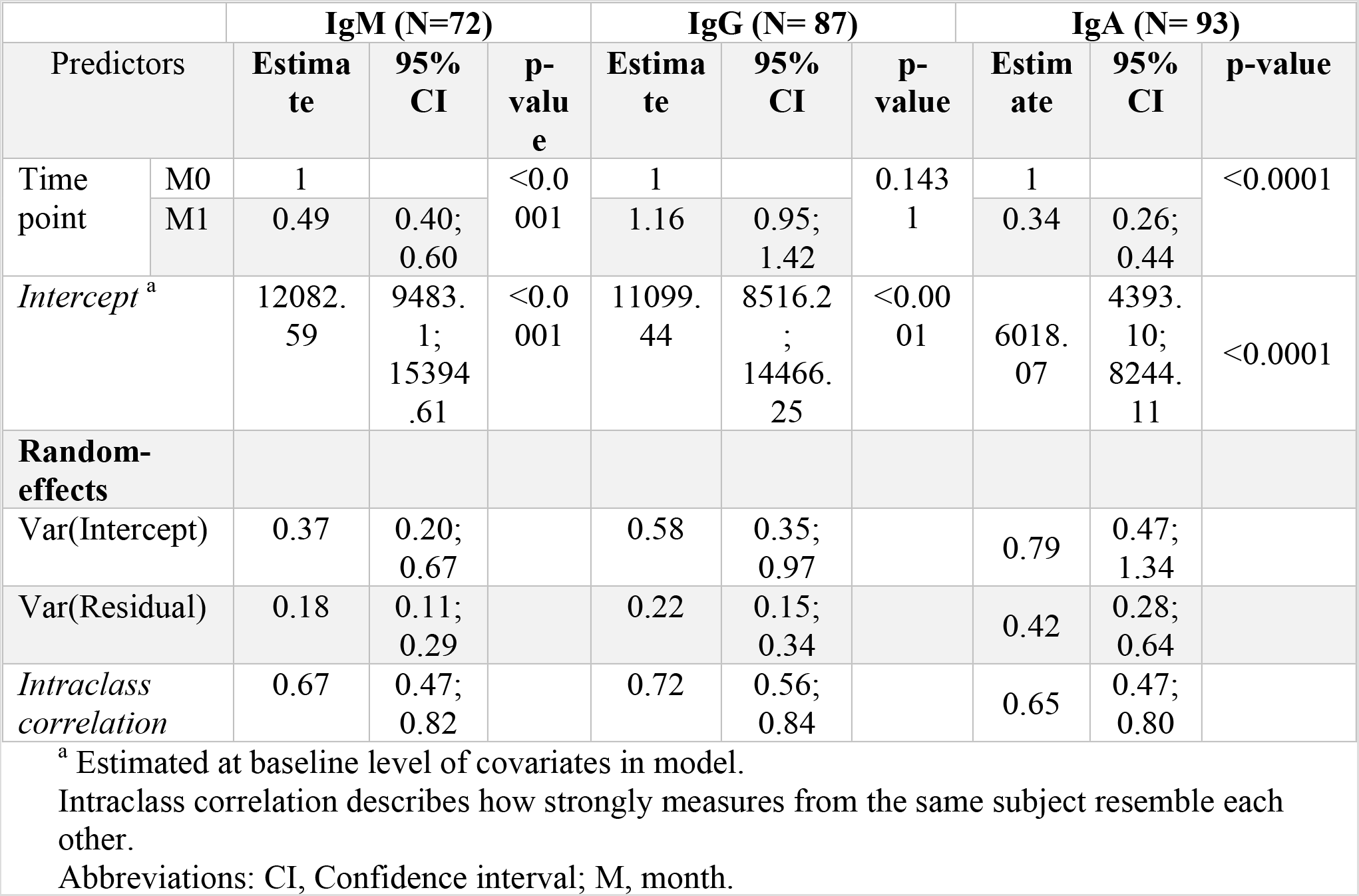
Rate of antibody decay calculated by mixed-effects linear models

**Figure 1.**
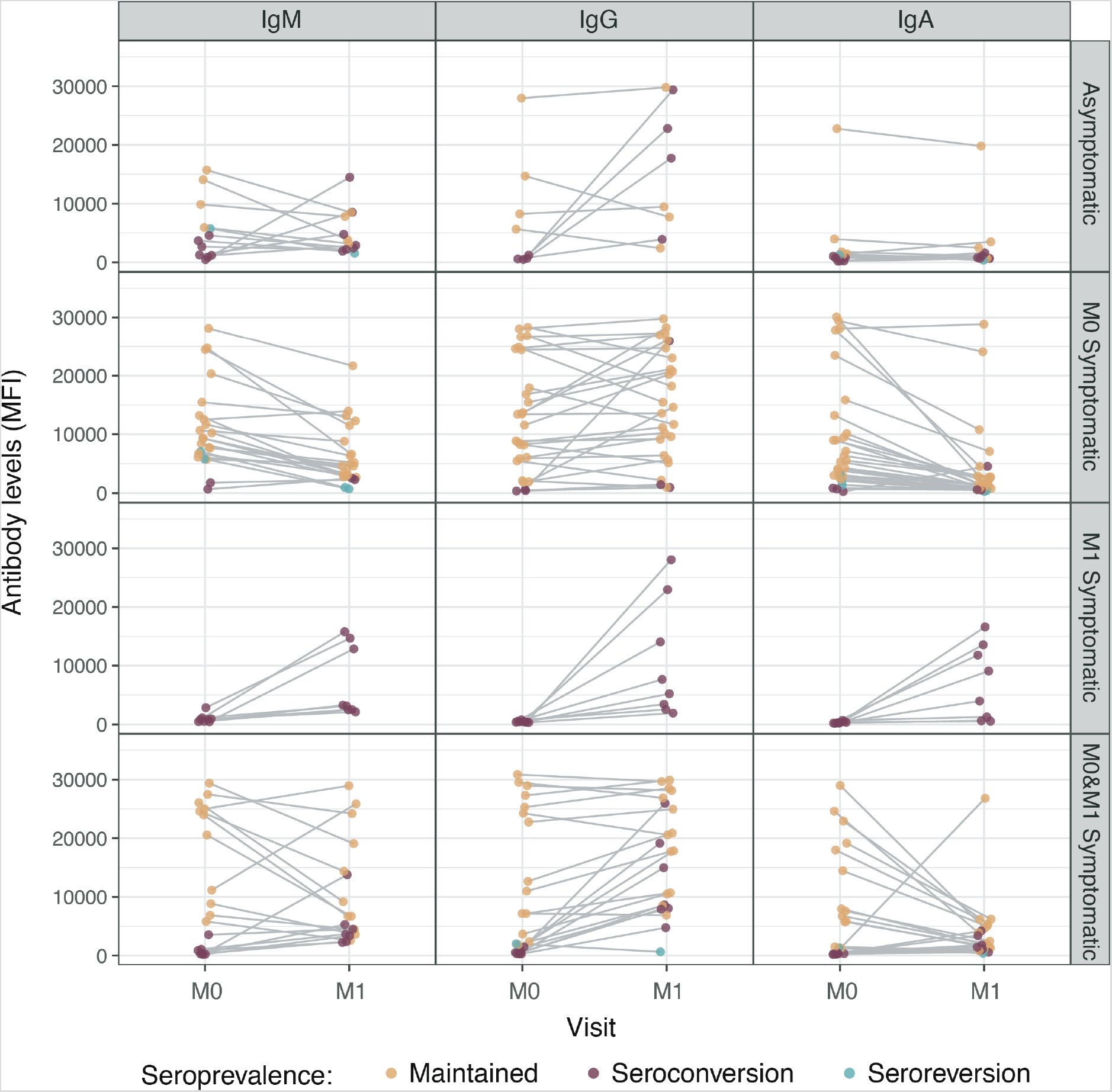
Kinetics in SARS-CoV-2 antibody levels in individuals seropositive at recruitment and/or at month 1. Levels (median fluorescence intensity, MFI) of IgM, IgG, and IgA against receptor-binding domain (RBD) of the SARS-CoV-2 spike glycoprotein stratified by asymptomatic participants and participants who reported COVID-19 compatible symptoms at recruitment (month 0, M0), month 1 (M1) or at both visits (M0&M1). Lines indicate paired samples. Yellow dots depict individuals who had detectable antibody levels at both study visits (IgM N = 33; IgG N = 42, IgA N = 41), burgundy and green dots show individuals who seroconverted for a particular isotype (IgM N = 24; IgG N = 22, IgA N = 24) and seroreverted (IgM N = 3; IgG N = 1, IgA N = 5), respectively, between visits.

IgA levels were higher in seropositive HCW reporting having had COVID-19 compatible symptoms (p< 0.001) and a similar trend was observed for IgM (p = 0.057, Figure 2). In addition, IgM levels were higher in those seropositive HCW with symptoms for > 10 days compared to seropositive HCW with shorter duration of symptoms, and a similar trend was observed for IgA (Figure 2). Age and sex were not associated with antibody levels (Figure 2 and Supplementary Figure 2).

**Figure 2.**
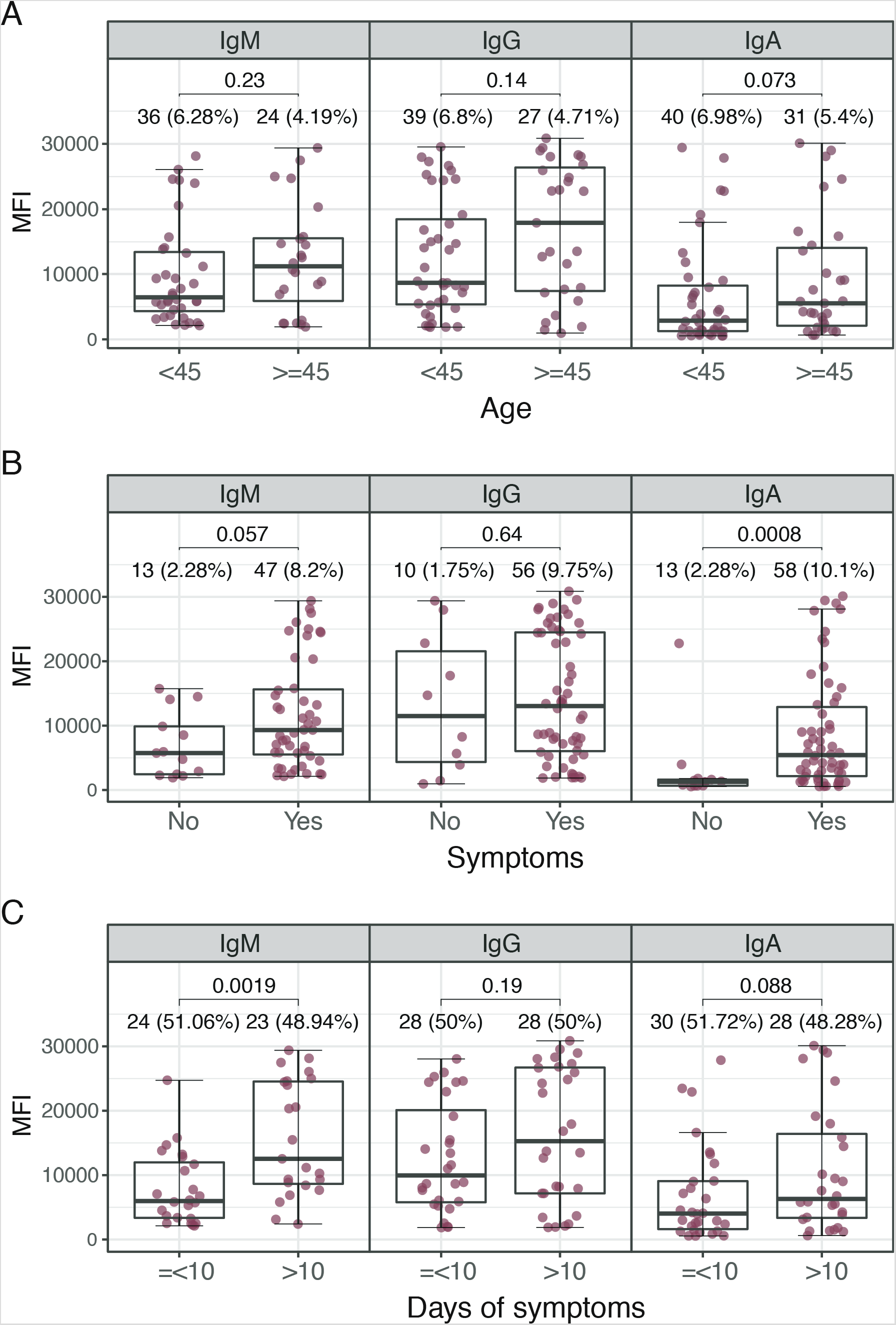
SARS-CoV-2 antibody levels by demographic and clinical variables. Levels (median fluorescence intensity, MFI) of IgM (N = 60), IgG (N = 66), and IgA (N = 71) against receptor-binding domain (RBD) of the SARS-CoV-2 spike glycoprotein stratified by (A) age, (B) presence of symptoms and (C) duration of symptoms. Graphs show data from accumulative month 0 and month 1 seropositive individuals: month 0 antibody levels from seropositive individuals at month 0 plus month1 antibody levels from individuals who seroconverted from month 0 to month 1. Percentages indicate the sum of proportions of seropositive subjects from recruitment and month 1 within each category of the x-axis with respect to the total number of samples from each visit (A and B) or the proportion of individuals within each category of the x-axis with respect to the total number of seropositive symptomatic (C). The center line of boxes depicts the median of MFIs; the lower and upper hinges correspond to the first and third quartiles; the distance between the first and third quartiles corresponds to the interquartile range (IQR); whiskers extend from the hinge to the highest or lowest value within 1.5 × IQR of the respective hinge. Wilcoxon rank test was used to assess statistically significant differences in antibody levels between groups.

Among HCW with positive rRT-PCR, IgM levels peaked around 18 days since the first positive rRT-PCR, declined during 30 days after the positive rRT-PCR and then seemed to stabilize (Figure 3A). IgA levels followed a similar pattern with a slightly earlier peak. Instead, IgG levels increased until 40 days since the first positive rRT-PCR and no decrease was observed thereafter. Similar kinetics were observed for antibody levels since onset of symptoms among seropositive HCW reporting having had symptoms (Figure 3B). However, antibodies peaked some days later compared to the kinetics by days since rRT-PCR, and IgM levels had a second lower peak around day 43.

**Figure 3.**
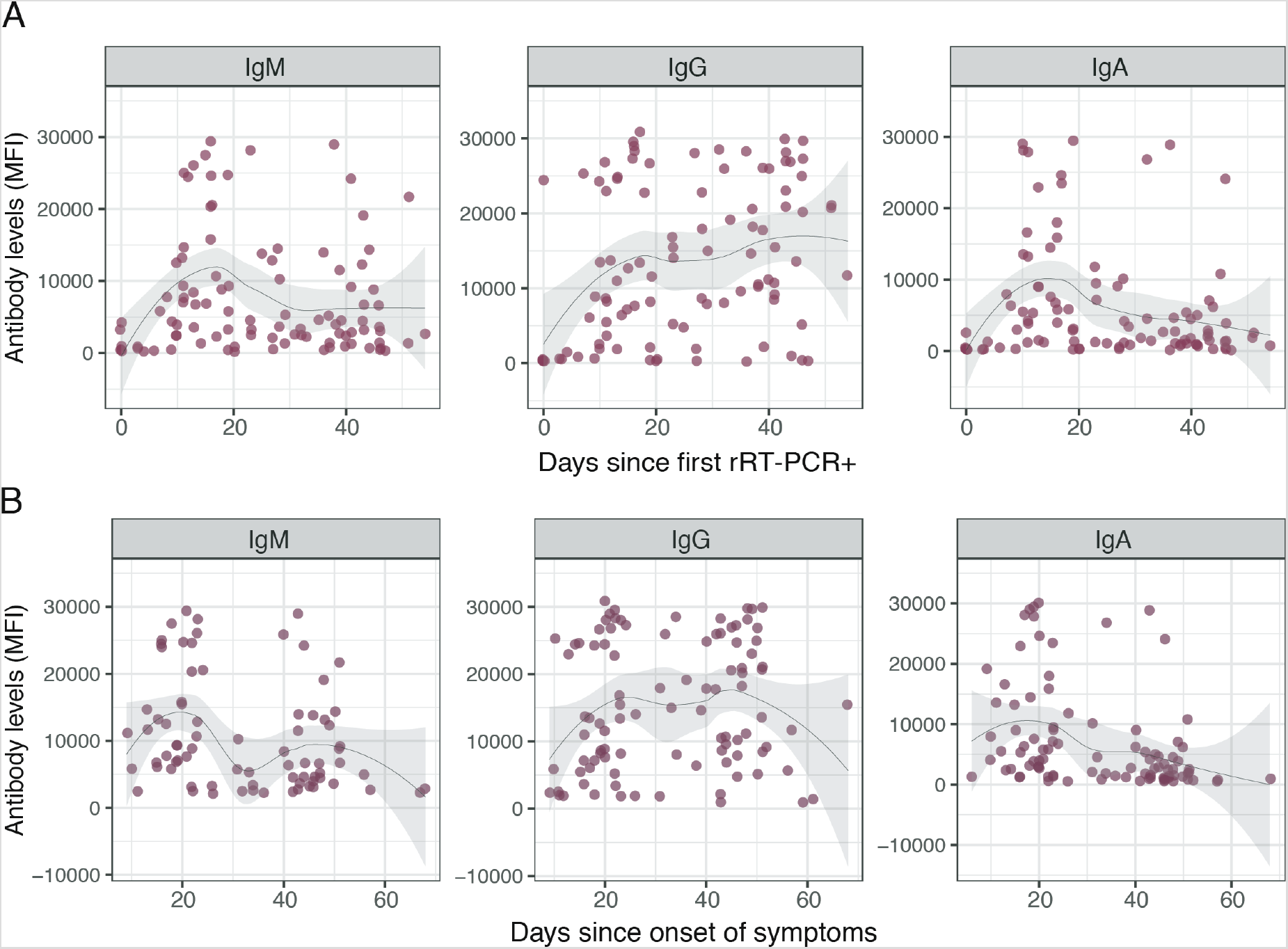
SARS-CoV-2 antibody levels by time since first rRT-PCR and onset of symptoms. Levels (median fluorescence intensity, MFI) of IgM, IgG, and IgA against receptor-binding domain (RBD) of the SARS-CoV-2 spike glycoprotein by (A) days since the first positive rRT-PCR, and (B) days since onset of any symptom. Graphs show pooled month 0 and month 1 data. Data in (A) are shown only for individuals with any rRT-PCR positive (N = 99). Data in (B) are shown only for seropositive individuals with any symptom compatible with COVID-19 (N = 77 for IgM, 96 for IgG, and 95for IgA). The fitting curve was calculated using the LOESS (locally estimated scatterplot smoothing) method. Shaded areas represent 95% confidence intervals.

### Antibody subclasses

IgG subclasses were measured only in IgG seropositive samples. IgG1 had the highest levels (and correlated with IgG), followed by IgG2, IgG3, and IgG4 (Figure 4A & B). Approximately 55% of the IgG positive samples had detectable IgG1 at month 0 and 1, and 60% and 79% had IgG2 at month 0 and 1, respectively (Figure 4C). Around 45% and 64% had detectable IgG3, and only 2% and 5% had IgG4 at month 0 and 1, respectively (Figure 4C). The levels of most subclasses were maintained or increased from month 0 to 1, and seroreversion was only observed for IgG2 and IgG4. The samples with the highest levels of IgG3 were IgG1 negative (Figure 4B).

**Figure 4.**
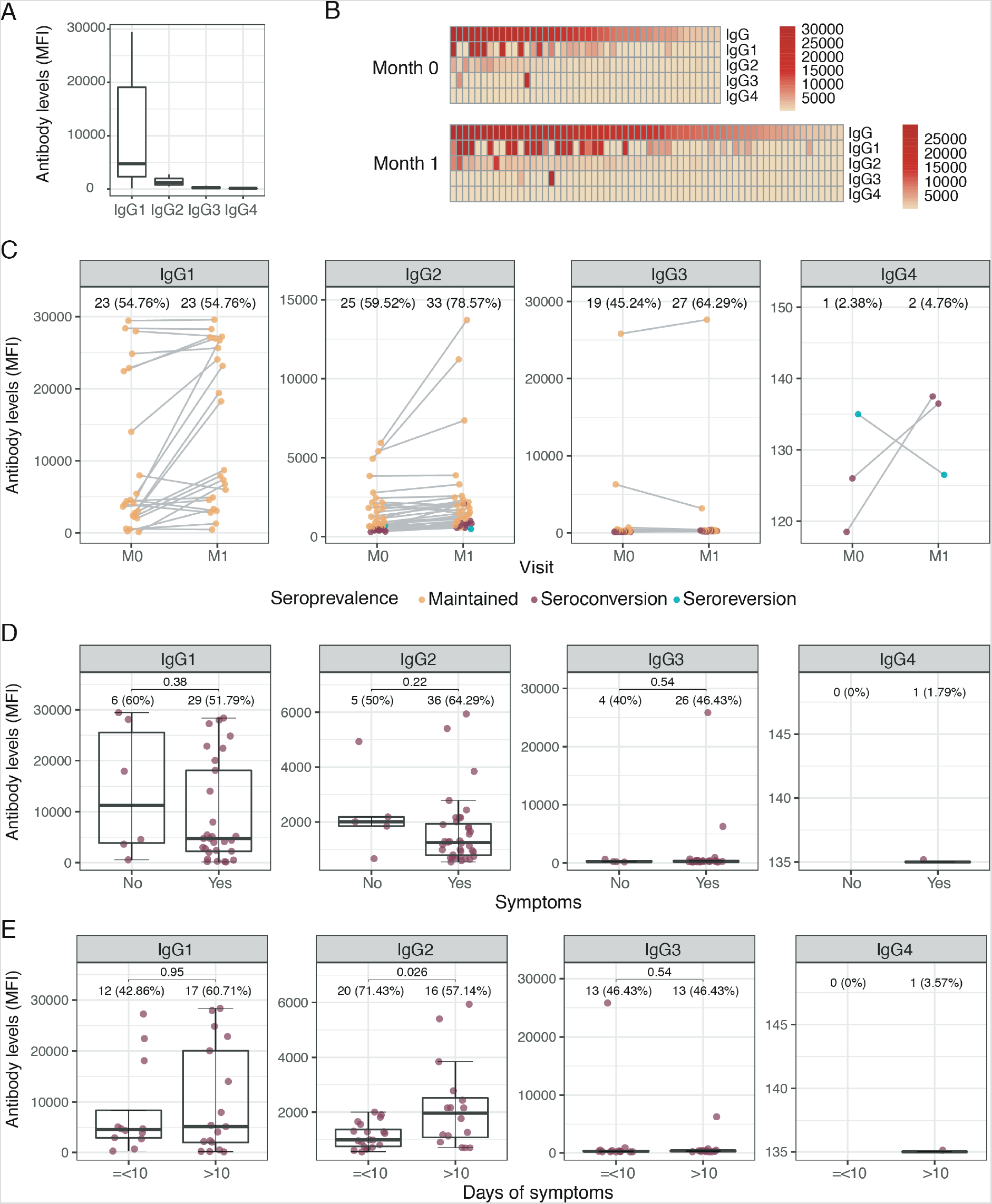
SARS-CoV-2 IgG subclass responses in IgG seropositive individuals. All panels show levels (median fluorescence intensity, MFI) of IgG1, IgG2, IgG3 and IgG4 against receptor-binding domain (RBD) of the SARS-CoV-2 spike glycoprotein. Y-axis scales are different for each subclass panel. (A) Boxplots (B) Heatmaps showing the MFI of IgG and IgG subclasses for all IgG seropositive individuals at recruitment and month 1 separately. (C) Kinetics of IgG subclass levels in seropositive individuals from M0 to M1. Lines indicate paired samples; yellow dots depict individuals who had detectable antibody levels at both study visits, burgundy and green dots show individuals who seroconverted and seroreverted, respectively, for the represented subclass between visits. (D) IgG subclass levels stratified by symptoms. (E) IgG subclass levels stratified by days of symptoms. (A), (D) & (E) show data from accumulative month 0 and month 1 seropositive individuals: month 0 antibody levels from seropositive individuals at month 0 plus month1 antibody levels from individuals who seroconverted from month 0 to month 1. Percentages indicate the proportion of seropositive subjects within each category of the x-axis. The center line of boxes depicts the median of MFIs; the lower and upper hinges correspond to the first and third quartiles; the distance between the first and third quartiles corresponds to the interquartile range (IQR); whiskers extend from the hinge to the highest or lowest value within 1.5 × IQR of the respective hinge. Wilcoxon rank test was used to assess statistically significant differences in antibody levels between groups.

IgG2 levels were higher in those IgG seropositive HCW who had more than 10 days of symptoms compared to those with lower duration of symptoms but no other associated factors were found (Figure 4D, Supplementary Figure 3).

Regarding IgA subclass, only IgA1 and no IgA2 was detected (Supplementary Figure 4).

## DISCUSSION

After one month of follow-up, we found a prevalence of 5% infections in HCW without a previous COVID-19 diagnosis or evidence of past infection. This is a substantial amount of new infections considering that (i) the accumulated prevalence of infection at recruitment was 11.2%, (ii) the peak of the pandemic had already passed, (iii) PPE were available, (iv) regular rRT-PCR screenings had been implemented for several weeks at the hospital, and (v) the population had been confined for almost 1.5 months. Interestingly, 64% of these infections were detected by serology only, probably reflecting infections occurring 1 to3 weeks before this survey.

The single factor showing the highest strength of association with newly detected past or present SARS-CoV-2 infection was having had any symptom compatible with COVID-19 in the previous month. Around 60% of the infected individuals were asymptomatic, which is a higher proportion than what we had previously reported at baseline [3], although in line with other studies reporting from 20 to 80% asymptomatic infections [16,17]. Consistently, we also found that working in a COVID-19 unit was not associated with SARS-CoV-2 infections [3] but, curiously, we found here that technicians had an increased risk. This may be due to a decreased perception of risk in this group in contrast to other job categories that may take more precautions due to their direct contact with COVID-19 patients.

Overall, IgM and IgA levels decreased substantially within a month and we estimated 2.77 and 2.26 months to IgM and IgA seroreversion, respectively. Of note, from the 54 seropositive HCW at recruitment, 8% had seroreverted for IgM and 11% for IgA. Time to seroreversion could not be calculated for IgG because many individuals had an increase in the antibody levels probably due to the short time since infection and the delayed peak response of IgG compared to IgA and IgM. The overall decrease of IgM and IgA but not IgG and the observed curves of antibody levels by days since the first positive RT-PCR are consistent with previously reported data [18,19] and with the expected patterns of an antibody response: IgM and IgA peak and then decline early after an infection and are typically short-term responses, while IgG peaks and decays later to a stable titer that is maintained over time. Also, IgG has a half-life of 21 days (less for IgG3) [20], whereas half-life of IgA and IgM is 4-9 days [21,22]. Nevertheless, emerging data indicate that SARS-CoV-2 IgG responses may wane quickly over time [23] to undetectable levels in a considerable proportion of individuals [24]. In our study, only 1 individual seroreverted for IgG. Antibody decay and seroreversion has enormous implications for the correct interpretation of serosurveys and could indicate waning protection and difficulties to achieve herd immunity.

We confirmed that IgA levels are higher in symptomatic individuals compared to the asymptomatic, and that IgM levels positively correlate with the duration of symptoms [3].

Despite not having found statistical differences for IgG levels, increasing evidence suggest that levels and duration of antibodies are also higher in symptomatic and in moderate-severe patients than mild cases [24,25]. In addition, we found 5 non-responders with more than 20 days since the first positive rRT-PCR. Lower antibody levels in asymptomatic and mild cases and antibody non-responders would also affect seroprevalence studies and could imply lower protection to reinfection, although infected individuals also mount T cell responses [26] which may independently protect from infection.

More than half of the seropositive HCW for IgG had IgG1 and IgG2 responses, whereas IgG3 responses were detected in less individuals. This finding differs from two other studies showing dominance of IgG3 and almost no IgG2 responses in COVID-19 patients [10,11]. IgG subclass responses increased from month 0 to month 1 in most of the individuals and, interestingly, many seroconverted during this month of follow-up for IgG2 and IgG3. Overall, antibody levels were higher for IgG1 than the other isotypes, following the relative abundance of these isotypes in plasma (IgG1 >IgG2 >IgG3 >IgG4). We did not find any factor associated with IgG subclass levels, with the exception of IgG2 levels positively correlating with higher duration of symptoms. Class-switch recombination to IgG2 occurs after IgG1 during the course of the immune response to limit inflammation [27]. While IgG1 are typically pro-inflammatory and have effector functions resulting in infection clearance through efficient binding to Fc receptors and complement, IgG2 has a decreased binding [27]. The association of IgG2 levels with duration of symptoms could reflect an anti-inflammatory response elicited by higher persistence of viruses and inflammation. Conversely, IgG2 could be contributing to persistence of symptoms through competition with IgG1, causing less efficient clearance of viruses.

The main limitation of this study is the small sample size for the analysis of factors associated with SARS-CoV-2 infection. In addition, there may be a recall bias in some reported data such as symptoms. In addition, antibody responses were only analyzed using one antigen and other viral proteins may elicit different responses in different populations [14], thus we could have slightly underestimated the overall seroprevalence of infection. Finally, kinetics of antibody responses and antibody decay rates have to be interpreted with caution as only two timepoints have been analyzed and rates may change depending on the baseline levels, and if levels are measured at the peak response or at the later steady-state period. Data from next timepoints will complete kinetics information.

Our findings reinforce the importance of strengthening SARS-CoV-2 surveillance among HCW. Despite having implemented regular rRT-PCR screenings, SARS-CoV-2 infections may go undetected. The lower antibody levels in the asymptomatic and mild cases, and the decay of IgA and IgM, have implications for seroprevalence studies as these isotypes may be undetectable 1-2 months post-infection. Although we could not show any evidence on IgG antibody decay after around 2 months from initial infection, we hope that subsequent surveys might provide some insight on this decay. Longer follow-up visits of this cohort will allow assessing the duration of IgG and IgG subclass responses and their role in protection from disease and reinfection.

## Data Availability

Data are provided in the manuscript and supplementary materials. Individual raw data are available upon request

## Acknowledgments

Special thanks to the HCW who participated in this study and all HCW who altruistically put their own lives at risk during the pandemic. We are grateful to F. Krammer for donation of RBD protein. We thank also Antoni Plasencia, Denise Naniche, Gonzalo Vicente, Caterina Guinovart, Martine Vrijheid, Matiana González, María Tusell, José Muñoz, Cristina Castellana, and administrative department (ISGlobal); Juan Valcárcel (CRG), Elías Campo (IDIBAPS), and nurses from Occupational Health and Preventive Medicine departments (HCB); Aida González (BETA Implants), Maria Jesús Mustieles, Jordi Vila and Mireia Navarro (HCB).

## SUPPLEMENTARY INFORMATION

### Supplementary Methods

#### Study design

Inclusion criteria included being an adult (>17 years) worker at HCB. Exclusion criteria included: (a) absenteeism from workplace in the last 30 days (i.e., on vacation, sick leave, sabbatical), (b) working exclusively outside the HCB or Maternity main buildings with no interaction with patients on a daily basis, (c) retirement or end-of-contract planned within one year after the recruitment date, and (d) participating in COVID-19 clinical trials for preventive or treatment therapies. Data included demographics, occupation, COVID-19 risk factors, clinical information related to COVID-19 compatible symptoms during the previous month, and history of rRT-PCR testing and contacts with cases.

#### SARS-CoV-2 rRT-PCR

RNA was extracted using the Quick-DNA/RNA Viral MagBead kit (Zymo) and the TECAN Dreamprep robot as described in the baseline manuscript [3]. Five microliters of RNA solution were used to amplify SARS-CoV-2 N1 and N2 regions, and the human RNase P gene as control, using probes, primers and cycling conditions described in the CDC-006-00019 CDC/DDID/NCIRD/ Division of Viral Diseases protocol (3/30/2020 release, Supplementary Note 1). Positive and negative controls were included in each batch of RNA extractions and rRT-PCR reactions [3]. A positive result was considered if the Ct values for N1, N2 and RNase P were below 40. Samples discordant for N1 and N2 were repeated, and samples with a Ct ≥ 40 for RNase P were considered as invalid.

##### Quantification of SARS-CoV-2 IgM, IgG and IgA by Luminex

RBD was coupled to magnetic microspheres from Luminex Corporation (Austin, TX). Antigen-coupled beads were added to a 96-well µClear^®^ flat bottom plate (Greiner Bio-One, 655096) at 2000 beads/well in a volume of 90 µL/well of phosphate buffered saline +1% bovine serum albumin + 0.05% sodium azide (PBS-BN). Next, 10 µl of test plasma samples (final dilution 1/500), 10 µl of a positive control (at four dilutions, 1/500, 1/2000, 1/8000 and 1/32000), 10 µl of two negative controls (final dilution 1/500), and 10 µl of PBS-BN as blank control were added per plate. Plates were incubated at room temperature (RT) for 2 h on a microplate shaker at 500 rpm and protected from light. Plates were washed three times with 300 µl/well of PBS-Tween20 0.05%, using a magnetic manual washer (Millipore, 43-285). A hundred microliters of biotinylated secondary antibody diluted in PBS-BN (anti-human IgG, B1140, 1/1250; anti-human IgM, B1265, 1/1000; or anti-human IgA, SAB3701227, 1/500; Sigma) were added to all wells and incubated for 45 min at 500 rpm at RT and protected from light. Plates were washed three times and 100 µL of streptavidin-R-phycoerythrin (Sigma, 42250) diluted 1:1000 in PBS-BN were added and incubated during 30 min at 500 rpm, RT and protected from light. Plates were washed three times, and beads resuspended in 100 µl of PBS-BN and kept overnight at 4 °C, protected from light. The next day, plates were read using a Luminex xMAP^®^ 100/200 analyzer with 70 µl of acquisition volume per well, DD gate 5000–25000 settings, and high PMT option. Crude median fluorescent intensities (MFI) and background fluorescence from blank wells were exported using the xPONENT software.

Sensitivity of the assay using samples from participants with positive SARS-CoV-2 rRT-PCR and with more than 10 days since the onset of symptoms was 97% for IgA and IgG and 75% for IgM, with specificities of 100% for IgG and IgM and 98% for IgA [3].

##### Quantification of SARS-CoV-2 IgG and IgA subclasses by Luminex

RBD was coupled to magnetic microspheres from Luminex Corporation (Austin, TX). Briefly, 2000 antigen-coupled microspheres in a volume of 50µL of PBS-BN were incubated with 50µL of test samples at the dilutions 1/100 and 1/500 during 1 hour on a microplate shaker at 500 rpm and protected from light. Plates were washed three times with 300 µl/well of PBS-Tween20 0.05%, using a magnetic manual washer (Millipore, 43-285). For IgG1 and IgG3, 100 µL of secondary antibody (anti-human IgG1-Biotin from Abcam ab9775 at 1/2000, and anti-human IgG3-Biotin from Sigma-Merck B3523 at 1/250) were added and incubated 45 min, followed by 3 washes and 100 µL of Streptavidin-R-Phycoerythrin (Sigma-Merck 42250 at 1/1000) incubated during 30 min at 500 rpm, RT and protected from light. For IgG2 and IgG4, 100 µL of secondary antibody (anti-human IgG2 at 1/500, and anti-human IgG4 at 1/500, from ThermoFisher, MA1-34655 and MA5-16716, respectively) were added and incubated 45 min, followed by 3 washes and 100 µL of a tertiary antibody (anti-mouse IgG-Biotin from Sigma-Merck B7401 at 1/40000 for IgG2, and 1/10000 for IgG4) incubated during 45 min at 500 rpm, RT and protected from light. After 3 washes 100uL of Streptavidin-RPhycoerythrin was added and plates were incubated 30 min at 500 rpm, RT and protected from light. For IgA1 and IgA2, 100 μL of secondary antibody-Phycoerythrin (mouse anti-human IgA1 RPE conjugate and IgA2 RPE conjugate at 1/200, from MOSS, 050520ZD05 and 050520ZD06, respectively) were added and incubated 30 min protected from light. Finally, after 3 washes, beads were resuspended in 100µl of PBS-BN and read using the Luminex xMAP^®^ 100/200 analyzer with 70 µl of acquisition volume per well, DD gate 5000–25000 settings, and high PMT option. At least 50 beads were acquired per sample. Crude median fluorescent intensity (MFI) and background fluorescence from blank wells were exported.

### Supplementary Tables

**Table S1.**
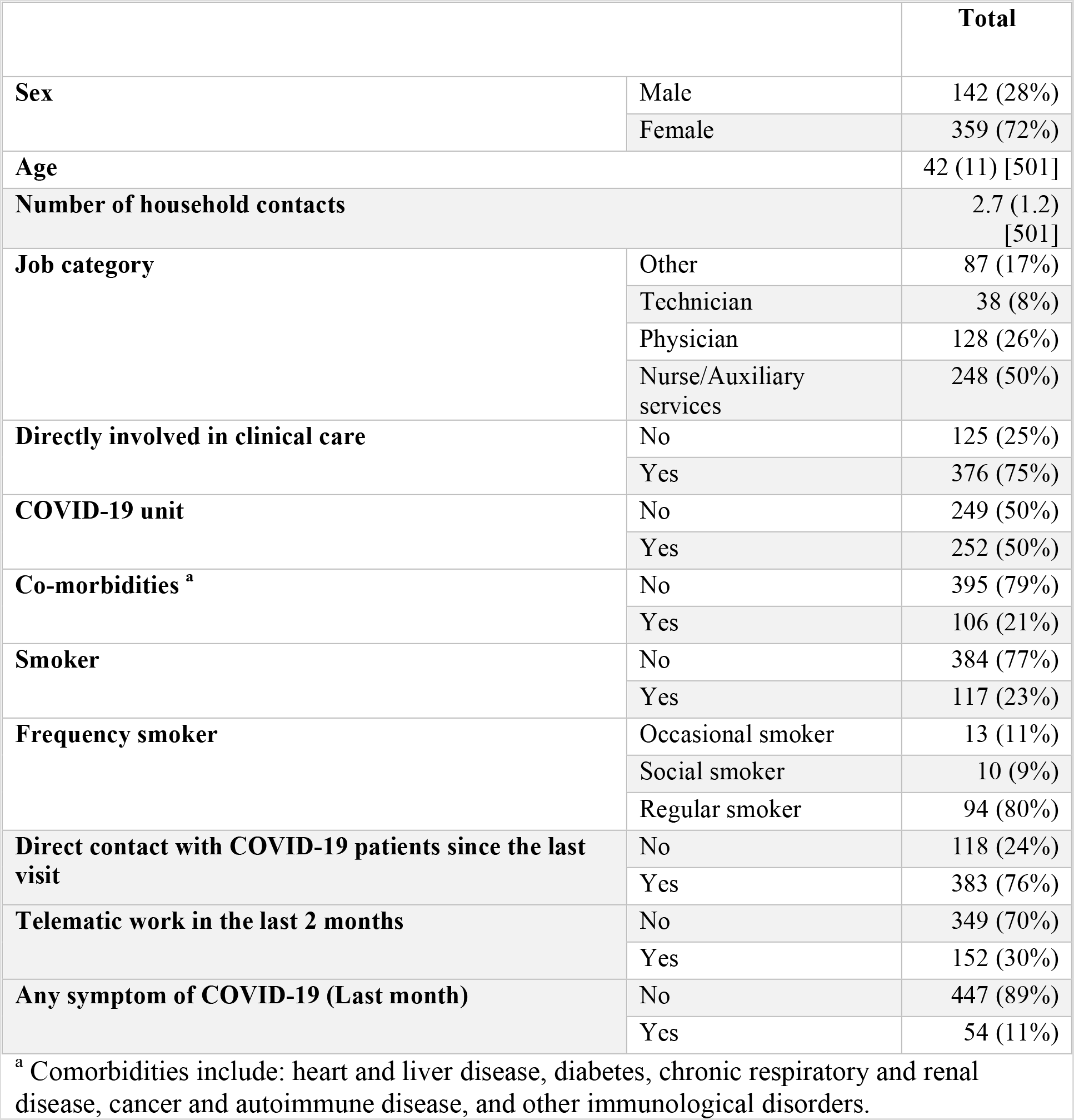
Characteristics of HCW without any evidence of current or past infection at baseline (month 0).

**Table S2.**
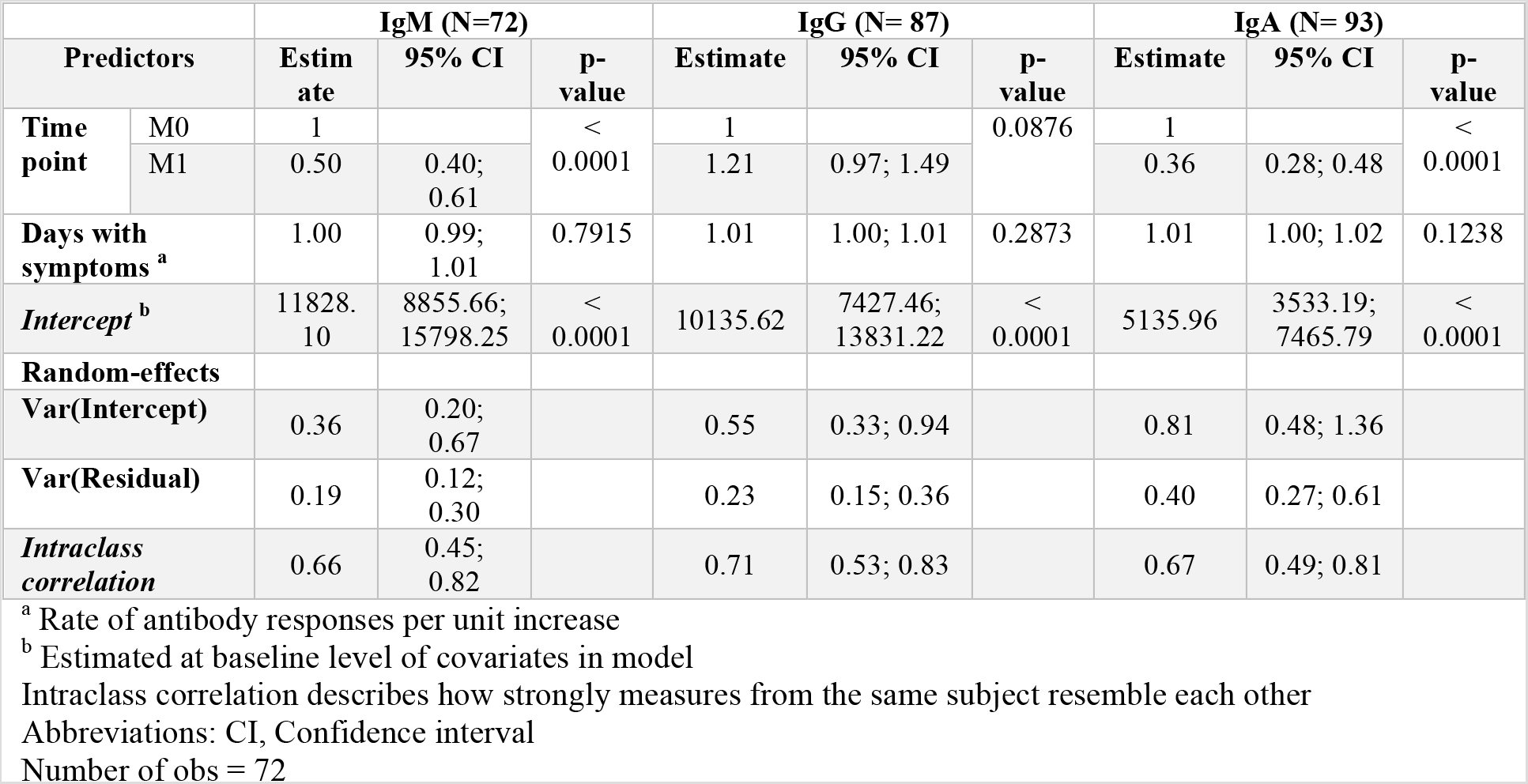
Rate of antibody decay calculated by mixed-effects linear models adjusted by days since onset of symptoms

### Supplementary Figures

**Figure S1.**
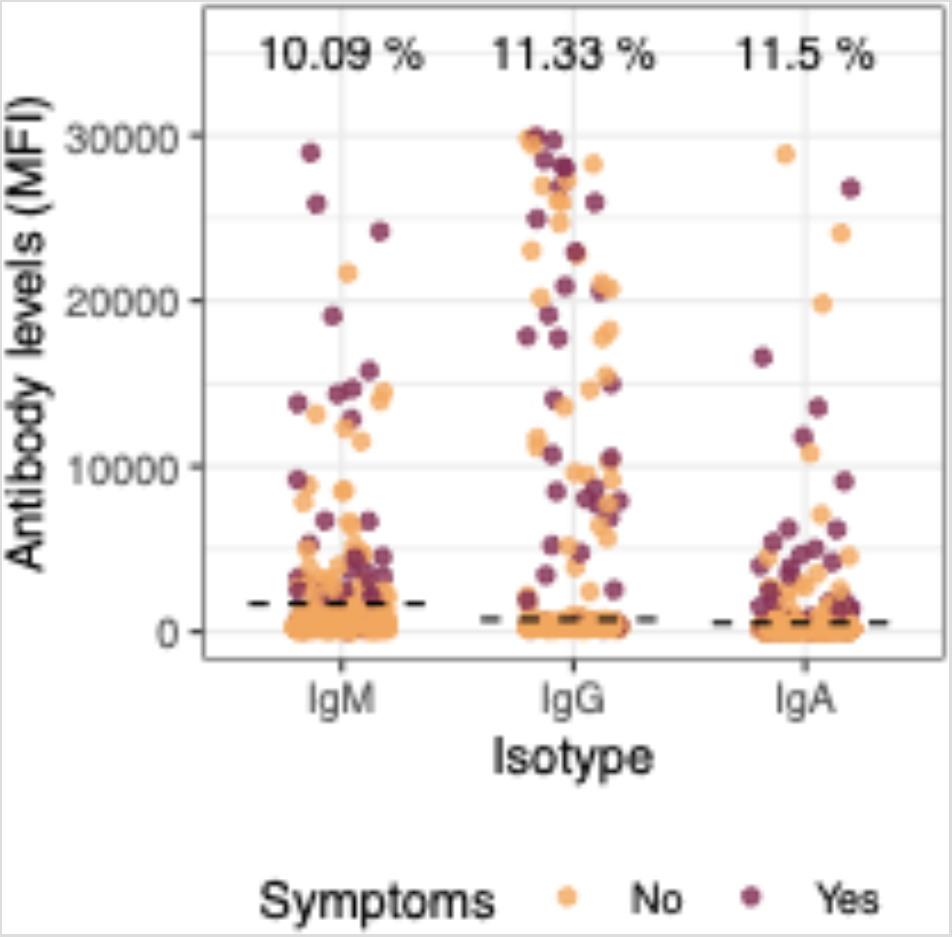
SARS-CoV-2 antibody levels (median fluorescence intensity, MFI) of IgM, IgG, and IgA against receptor-binding domain (RBD) of the SARS-CoV-2 spike glycoprotein in all study participants at month 1. Percentages indicate the proportion of seropositive individuals within each immunoglobulin. Dashed line represents the seropositivity cutoff calculated as 10 to the mean plus 3 standard deviations of log10-transformed MFIs of 47 prepandemic negative controls. Burgundy dots represent symptomatic individuals and yellow dots asymptomatic participants.

**Figure S2.**
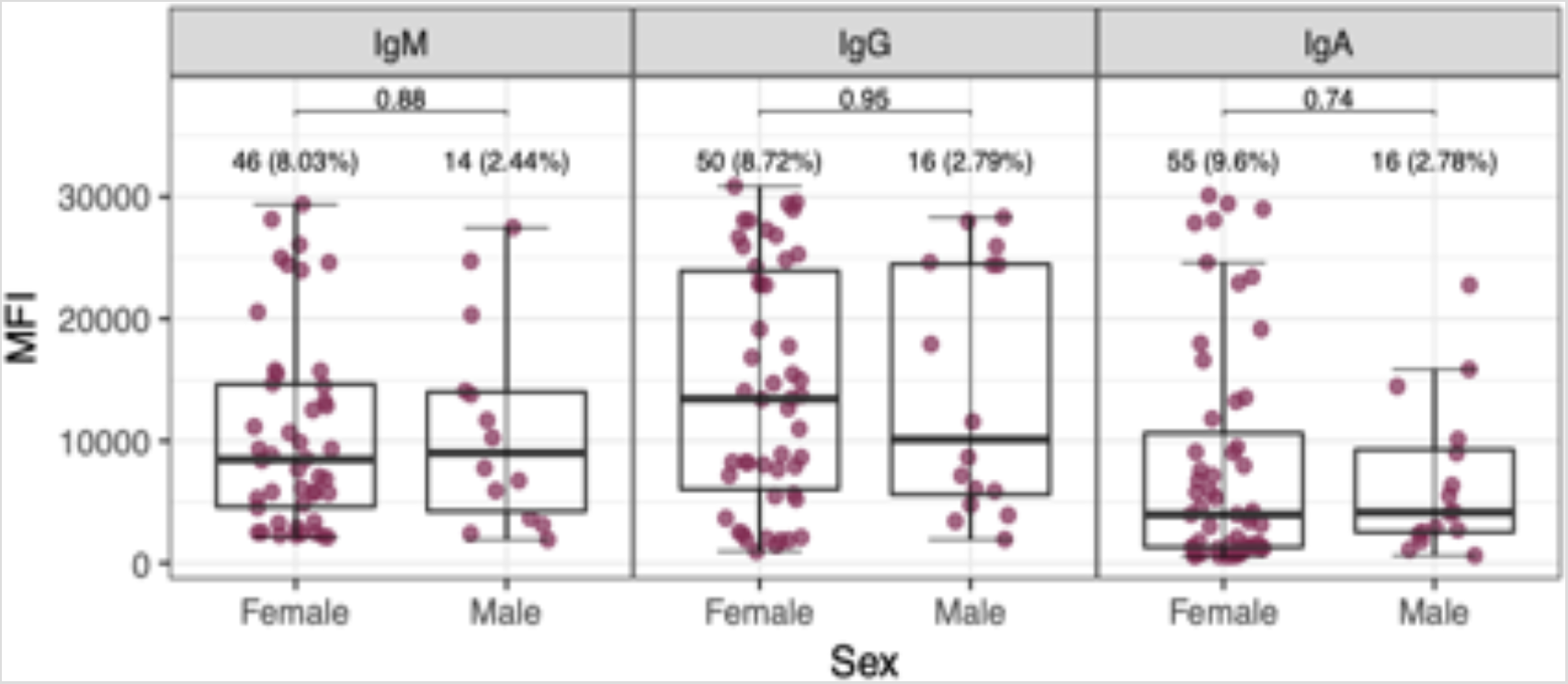
SARS-CoV-2 antibody levels by sex. Levels (median fluorescence intensity, MFI) of IgM (N = 60), IgG (N = 66), and IgA (N = 71) against receptor-binding domain (RBD) of the SARS-CoV-2 spike glycoprotein. Graphs show data from accumulative month 0 and month 1 seropositive individuals: month 0 antibody levels from seropositive individuals at month 0 plus month1 antibody levels from individuals who seroconverted from month 0 to month 1. Percentages indicate the proportion of seropositive subjects within each category of the x-axis with respect to the total number of samples from each visit. The center line of boxes depicts the median of MFIs; the lower and upper hinges correspond to the first and third quartiles; the distance between the first and third quartiles corresponds to the interquartile range (IQR); whiskers extend from the hinge to the highest or lowest value within 1.5 × IQR of the respective hinge. Wilcoxon rank test was used to assess statistically significant differences in antibody levels between groups.

**Figure S3.**
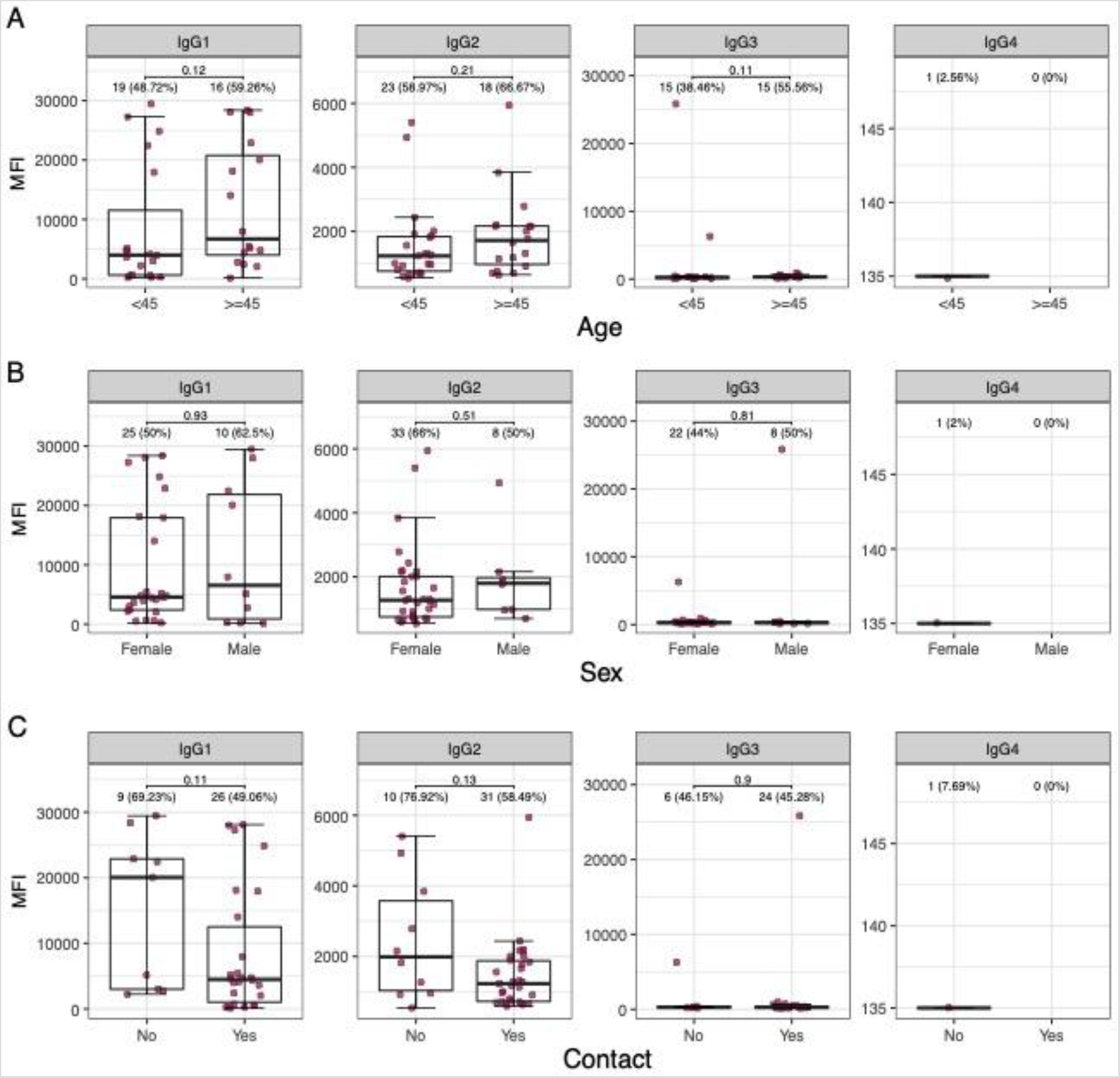
Levels of SARS-CoV-2 IgG subclasses in IgG seropositive individuals by demographic and clinical factors. SARS-CoV-2 antibody levels by demographic and clinical variables. Levels (median fluorescence intensity, MFI) of IgG, IgG2, IgG3 and IgG4 against receptor-binding domain (RBD) of the SARS-CoV-2 spike glycoprotein stratified by (A) age, (B) sex, and (C) known contact with COVID-19 cases. Graphs show data from accumulative month 0 and month 1 seropositive individuals: month 0 antibody levels from seropositive individuals at month 0 plus month1 antibody levels from individuals who seroconverted from month 0 to month 1. Percentages indicate the proportion of seropositive subjects within each category of the x-axis. The center line of boxes depicts the median of MFIs; the lower and upper hinges correspond to the first and third quartiles; the distance between the first and third quartiles corresponds to the interquartile range (IQR); whiskers extend from the hinge to the highest or lowest value within 1.5 × IQR of the respective hinge. Wilcoxon rank test was used to assess statistically significant differences in antibody levels between groups.

**Figure S4.**
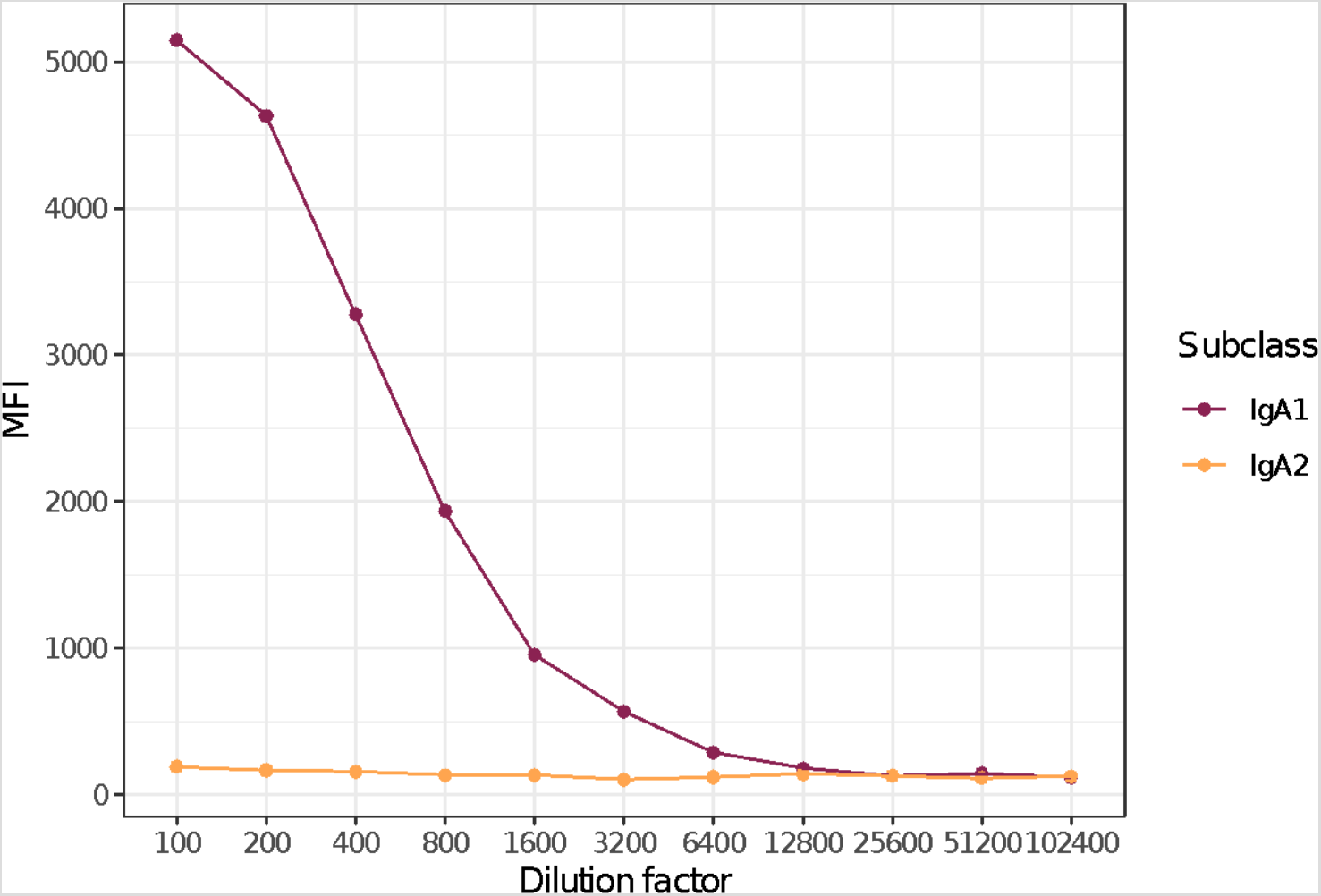
SARS-CoV-2 IgA subclass levels in a pool of samples from seropositive individuals. Levels (median fluorescence intensity, MFI) of IgA1 and IgA2 against receptor-binding domain (RBD) of the SARS-CoV-2 spike glycoprotein in serial dilutions of a pool of seropositive samples.

